# A putative hepatitis B virus sequence motif associated with hepatocellular carcinoma in South African adults

**DOI:** 10.1101/2024.01.13.24301263

**Authors:** Tongai G Maponga, Anna L McNaughton, Cori Campbell, Mariateresa de Cesare, Jolynne Mokaya, Sheila F Lumley, David Bonsall, Camilla LC Ip, Haiting Chai, Christo Van Rensburg, Richard H Glashoff, Elizabeth Waddilove, Wolfgang Preiser, Jason T Blackard, M Azim Ansari, Anna Kramvis, Monique I Andersson, Philippa C Matthews

## Abstract

**Aim:** Chronic hepatitis B virus (HBV) infection is a major risk factor for hepatocellular carcinoma (HCC) particularly in African populations, in whom malignancy frequently presents at an advanced stage with poor outcomes. We derived HBV whole genome sequences (WGS) from individuals with HCC and compared them to sequences from individuals without HCC. **Methods:** We identified adults with HBV infection, with and without complicating HCC, in Cape Town, South Africa and utilized pan-genotypic probe-based enrichment followed by Illumina sequencing to derive HBV WGS.

**Results:** Compared to the non-HCC group, HCC patients were more likely to be male (p < 0.0001), older (p = 0.01), HIV-negative (p = 0.006), and to have higher HBV viral loads (p < 0.0001). Among 19 HCC and 12 non-HCC patients, genotype A dominated (74%), of which 96% were subtype A1. PreS2 deletions (Δ38–55) were enriched in HBV sequences from HCC patients (n = 7). The sequence motif most strongly associated with HCC comprised either deletion or polymorphism at site T53 in PreS2 – collectively coined ‘non-T53’ – together with a basal core promoter (BCP) mutation G1764A (AUROC 0.79).

**Conclusions:** In this setting, HBV sequence polymorphisms and deletions are associated with HCC, and ‘non-T53 + G1764A’ represents a putative signature motif for HCC. Additional investigations are needed to disaggregate the impact of age, sex and HIV status, to ascertain the extent to which viral polymorphisms contribute to oncogenesis, and to determine whether HBV sequence is a useful biomarker for risk stratification.

## INTRODUCTION

A high incidence of hepatocellular carcinoma (HCC) in the World Health Organization (WHO) African region (63,000 cases per annum in 2018) [1,2] reflects the prevalence and distribution of hepatitis B virus (HBV) infection [3,4]. The substantial individual and societal impact of HBV- associated HCC in African populations is related to disease affecting young adults and presenting with advanced malignancy, leading to high mortality [5,6]. International targets aim to eliminate the public health threat of HBV infection, with HCC as an important area of focus [7].

In Southern Africa, infection with genotype-A HBV has a strong association with HCC [8,9]. Malignant transformation can occur as a result of chronic inflammatory/fibrotic liver disease, HBV DNA integration into the host genome, cell stress caused by accumulation of aberrant viral proteins [10], and/or a direct influence of HBV genes (particularly HBx) [11]. Specific viral polymorphisms have been associated with HCC, including truncated genes, pre-core insertions/deletions (‘indels’), and basal core promoter (BCP) mutations [12–14]. Such sequence changes can be used potentially to infer disease risk or prognosis [15–17]. However, further work is needed to better describe the mutational landscape of HBV in diverse viral genotypes, advance insights into specific associations between viral sequence polymorphisms and HCC, and determine their mechanistic impact. Evaluation is needed to determine whether viral sequence can inform clinical risk assessment, surveillance or interventions.

We undertook HBV sequence analysis from South African adults to explore viral sequence polymorphisms in those with and without HCC.

## METHODS

### Study samples

We retrospectively drew on banked serum samples from adults with a confirmed diagnosis of chronic HBV infection, in cohorts with and without HCC (described in [18] and [19] respectively) (**Suppl methods 1; Suppl Fig 1**). Ethical approval was granted from the health research ethics committee at the University of Stellenbosch (S13/04/072 and N11/09/284).

### Illumina sequencing, HBV genome assembly and phylogenetic analysis

We generated HBV sequence data based on an adapted version of a published Illumina protocol [20] (**Suppl methods 2**). HBV reads were mapped to reference sequences (genotypes A-I) prior to generating consensus WGS. A standard HBV reference strain (Genbank accession X02763, genotype A) was used for numbering positions in the genome.

For phylogenetic analysis, we aligned nucleotide alignments using new sequence data from this cohort, 61 full-length South African sequences from Genbank, and genotype reference sequences[21] using Clustal X 2.1 [22], and performed phylogenetic inference using a Bayesian Markov Chain Monte Carlo (MCMC) approach as implemented in the Bayesian Evolutionary Analysis by Sampling Trees (BEAST) version 1.10.4 program [2] (**Suppl methods 3**).

### Analysis

We identified HBV polymorphisms that have been previously associated with HCC [11,23–26] (**Table 1**), and explored the association between polymorphisms and HCC using area under the receiver-operating curve (AUROC). The frequency of deletions at each site was compared between HCC and non-HCC sequences. Statistical analyses were performed using GraphPad Prism v10 and STATA v17.0.

**Table 1:**
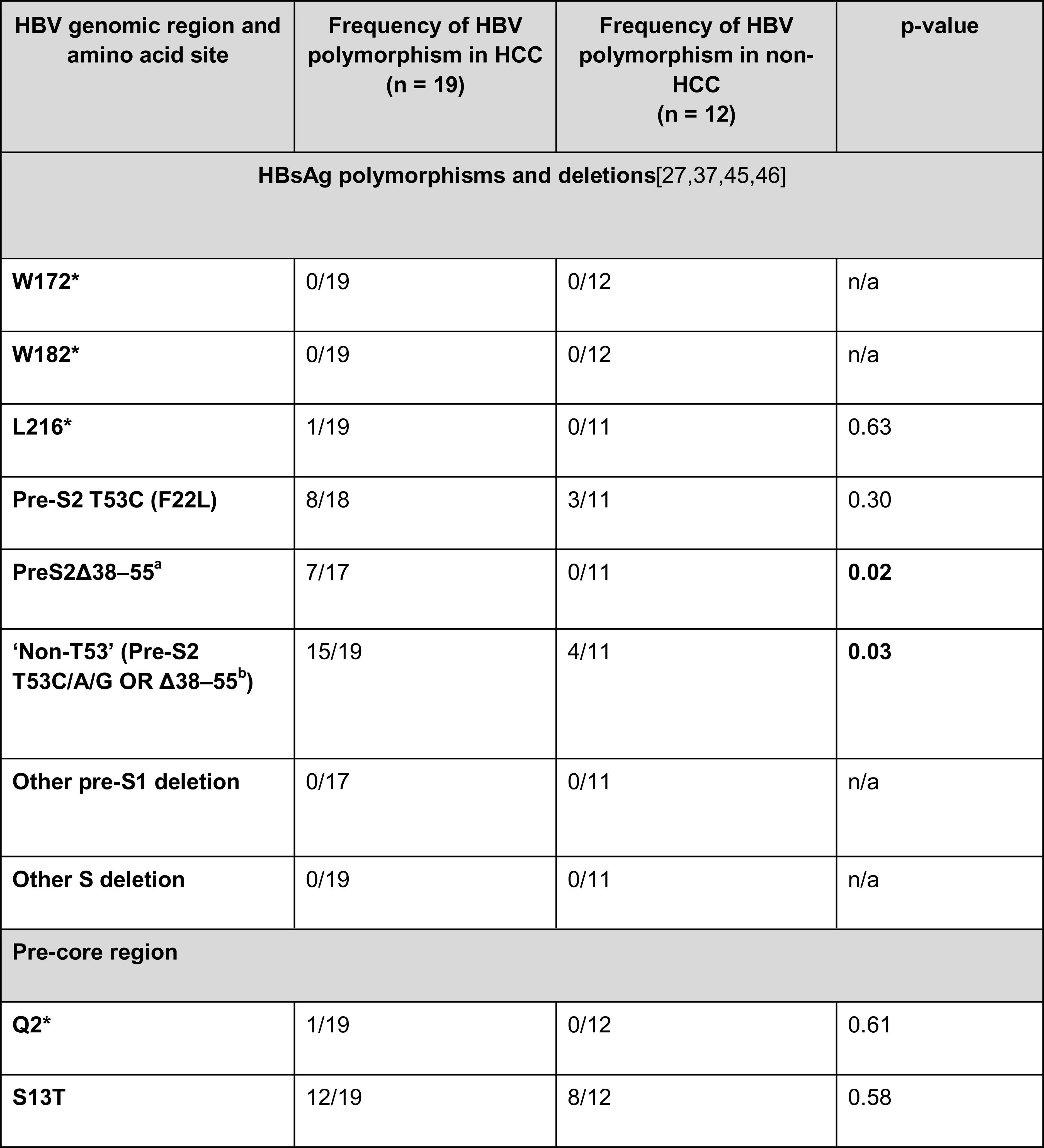
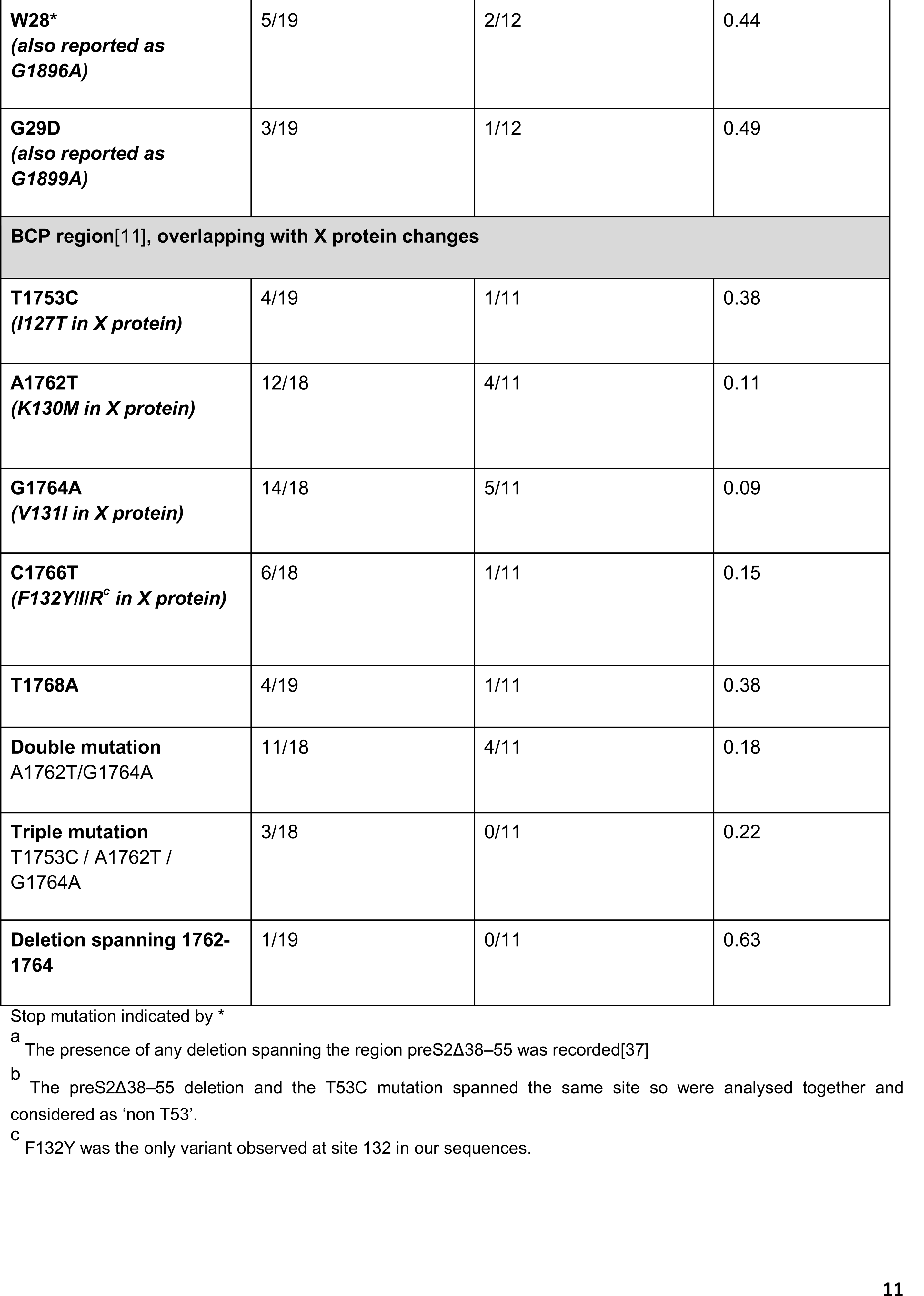
Frequency of HBV polymorphisms previously associated with hepatocellular carcinoma (HCC) identified in consensus sequences derived by Illumina among patients with and without a diagnosis of HCC. Frequencies are based on analysis of consensus sequences. Partial genomes were derived in some cases; hence, the denominator at certain sites is lower than the total number of samples sequenced.

## RESULTS

### HBV-associated HCC associated with male sex and higher HBV viral loads

We identified samples from 161 adults with chronic HBV infection of whom 68 had HCC and 93 not. Compared to the non-HCC group, those with HCC were more likely to be male (81% vs. 46%, respectively, p < 0.0001), older (median 41 vs. 36 years, p = 0.01), and had higher HBV DNA viral loads (median 5.2 vs 3.5 log_10_ IU/mL, p < 0.0001). HIV coinfection was present in 18 of 65 HCC cases (3 had no documented test result) and 46 of 93 non-HCC cases (27.8% vs 49.5%; p = 0.006) (**Suppl Table 1**).

We were able to undertake WGS HBV sequencing from 19 samples from the HCC group and 14 from the non-HCC group (**Suppl Methods 2**), with sufficient reads to construct WGS assemblies from 31 of these (19 HCC and 12 non-HCC (**Table 1; Suppl Fig 2**)). Sequence data can be accessed in GenBank project PRJEB71107.

Overall, genotype A predominated, accounting for 23 of 31 (74.2%), of which 22 were A1. There was no difference in genotype distribution between HCC and non-HCC groups (**Suppl Fig 1; Suppl Tables 2 and 3; Suppl Results 1**). HBV sequences from HCC and non-HCC groups were interspersed with other South African HBV sequences, suggesting there was no particular viral lineage associated with the development of HCC (**Suppl Fig 3**).

### Deletions and substitutions in HBV Pre-S2 significantly enriched in HCC

We examined HBV sequences for the presence of polymorphisms and deletions previously associated with HCC (**Table 1**). There was sequence coverage in the PreS2 region in 17 of 19 HCC cases, among which 7 of 17 had PreS2 Δ38–55 deletions, compared to none in the non-HCC group (p = 0.02) (**Figure 1A, 1B**). In the consensus sequences, deletions were observed ranging from 3 bp to 42 bp in length, with a mean of 21 bp. Start locations of the sequences varied, but all deletions terminated by nucleotide (nt) 55. We also evaluated the T53C substitution that has been reported in association with HCC[27]. In the HCC group, wild-type T53 was uncommon, due to a combination of deletions (n = 6), and substitutions (n = 9; T53C substitution in 8 and T53G in one). Thus, overall, the ‘non-T53’ motif occurred in 15/19 samples in the HCC group compared to 4/11 in the non-HCC group (p = 0.027; **Table 1**).

**Figure 1.**
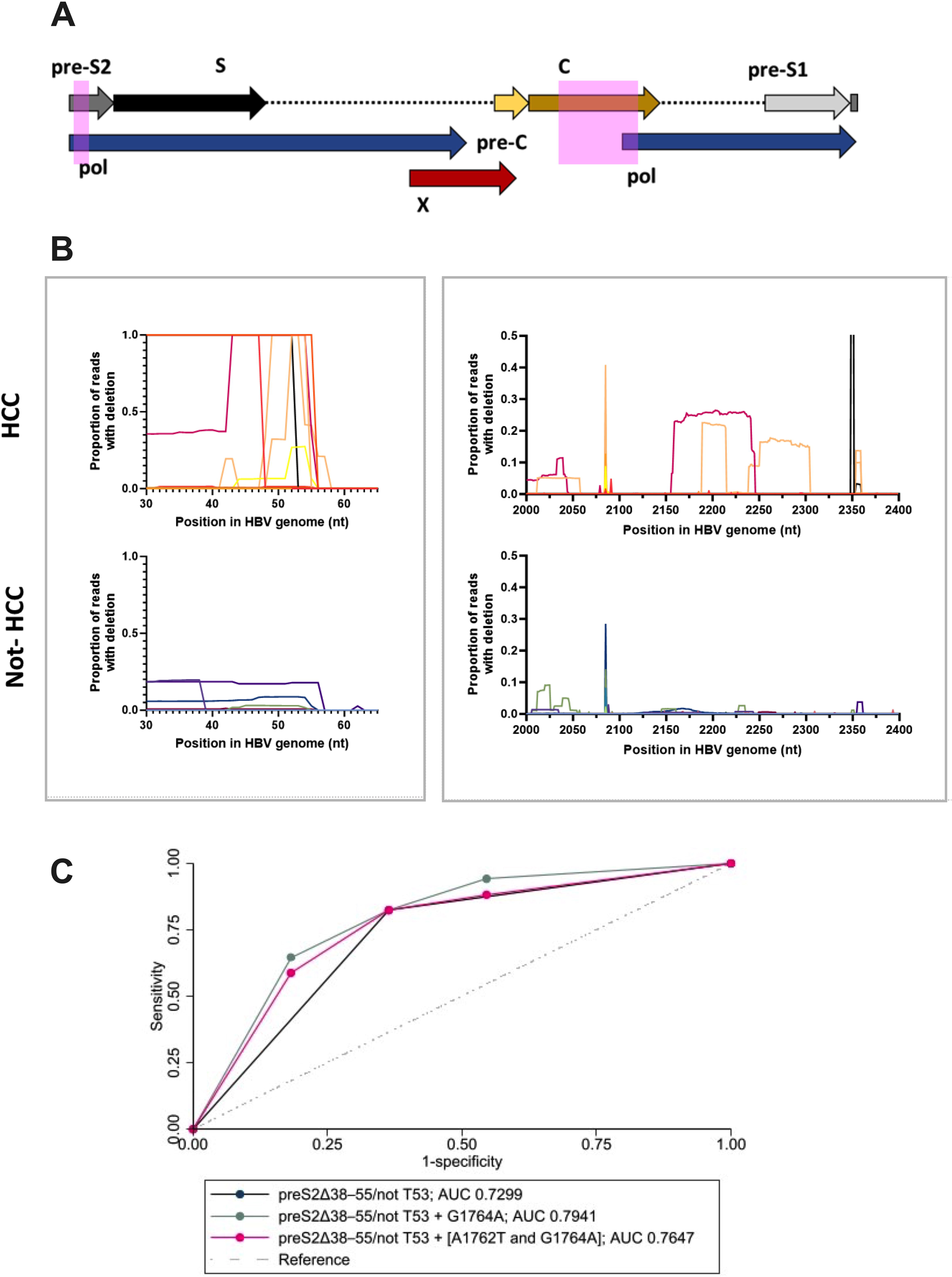
Legend: HBV Sequence motifs associated with HCC in South African adults. A: Schematic of HBV genome. Region of PreS2 and BCP polymorphisms are shown in pink highlights. B: Regions of the HBV genome showing the proportion of reads with deletions in at each site. Data are presented for individuals with HCC (top) and without HCC (bottom). Termination of deletions in 7 HBV sequences from the HCC group was at nt 52 (n = 1), nt 53 (n = 2), nt 54 (n = 2), and nt 55 (n = 2). Additional discussion of the minority variant deletions is provided in **Suppl results 2**. The bioinformatic pipeline did not provide deletion frequencies for reads when a deletion was the consensus at that site (the site was absent), so these sites have been assigned a frequency of 100% to generate these plots. C: Receiver operating characteristic (ROC) curves for pre-S2 deletions and T53 polymorphisms combined with BCP polymorphisms as a predictor of HCC status in HBV sequences. Curves are shown for combinations of PreS2 ‘not T53’ alone, or combined with the BCP polymorphisms G1764A and the double mutation A1764T /G1764A. Two sequences, HCC-34 and nHCC-12 were excluded from the analysis as there was no sequence for the region under consideration. AUC, area under the curve.

### BCP mutations

Mutations in the BCP region associated with HCC by other studies (A1762T, G1764A and the combination of both together) were more common in the HCC group; however, this was not significantly different from the non-HCC group (**Table 1**).

### Associations of combined polymorphisms with HCC

Finally, we analysed ‘non-T53’ together with the most common BCP mutation (G1764A) or the BCP double mutation (A1762T + G1764A). The combination of ‘non-T53’ with the G1764A mutation was the most strongly associated with HCC (AUROC = 0.79) (**Table 1**, **Figure 1C**).

## DISCUSSION

In this small sample set, HBV PreS2 ‘non-T53’ (either as a result of a mutation or deletion) in combination with the BCP G1764A polymorphism, is the sequence motif most strongly associated with HCC. Both of these sequence changes have been reported independently in South African sequence data [28–32]. However, to the best of our knowledge, this combined motif has not been previously studied. Further data are needed to evaluate the sensitivity and specificity of the association between this motif and HCC, and to determine the impact of potential confounders such as age, sex, HBV VL, HIV status, and antiviral treatment, which was not possible in this small cohort. Furthermore, there may be an impact of additional contributors which were not measured, such as host genetic/epigenetic factors, and external environmental factors [33–36]. As sequencing only generates data for high VL samples, there may be artificial enrichment for A1762T and G1764A mutations.

PreS2 deletions have also been reported in association with HCC in West African sequence data, potentially associated with aflatoxin exposure and/or conferring a viral fitness advantage[37]). Mechanistically, ‘non-T53’ may have an influence on PreS2 function or regulation and/or impact the spacer domain of the polymerase protein. Proposed oncogenic mechanisms include the accumulation of defective proteins in the endoplasmic reticulum (ER), stress responses resulting in DNA damage, centrosome over-duplication, and genomic instability [25,38–40]. PreS2 deletions start at a wide range of sites (sometimes prior to nt 38) but consistently terminate immediately prior to nt 55, where there is a highly conserved region of approximately 20 bp, and the first of several cysteine residues within a putative zinc finger domain, considered to be essential for reverse transcriptase activity [41]. Therefore, deletions downstream of nt 55 are likely to be detrimental to viral replication, potentially explaining why they all terminate within the same region.

A longer duration of infection may be needed for cumulative BCP mutations to develop, and an increased frequency of these polymorphisms has been reported as HBV infection progresses [42,43]. The increase in A-T-rich regions in the BCP region (nt 1762-1770) may be associated with upregulating viral transcription [44], and these mutations may influence malignant transformation mediated through the overlapping X gene [16].

In future, larger studies with matched case:control recruitment would allow multivariable analysis, investigation of the impact of sequence changes in diverse genotypes, and an unbiased, sequence agnostic approach to WGS changes (instead of focusing only on polymorphisms that have been previously reported). To date, sequencing protocols have been limited by high VL thresholds and/or cost. As the sensitivity of sequencing methods improves, it may be possible to sequence HBV genomes at lower VL, to analyse within-host sequence diversity, and to generate long-read sequencing[20] to support WGS haplotype analysis. We highlight the pressing need for the field to be expanded by the open-source sharing of HBV sequences together with clinical metadata.

In conclusion, we propose a novel HBV sequence motif that may be associated with HCC, but further work is required to determine the cause, effect and chronology of these sequence changes relative to clinical and demographic characteristics, and the evolution of HCC.

## Data Availability

Consensus sequences are available on line at https://www.doi.org/10.6084/m9.figshare.24874557.Other data produced in the present study are available upon reasonable request to the authors.

https://www.doi.org/10.6084/m9.figshare.24874557

## ACKNOWLEDGEMENTS

We are grateful to the staff and patients at our centres of recruitment for their support and participation.

## AUTHORS’ CONTRIBUTIONS

Concept: TGM, ALM, PCM

Patient recruitment: TGM, CVR, MIA

Ethics and governance: TGM, EW, MIA, RG Laboratory work: TGM, ALM, MdC, DB

Data analysis and curation: ALM, CLCI, MAA, CC, JM, JTB, DB, HC Wrote the original manuscript: ALM, CLCI, PCM

Edited and approved the manuscript: All authors Supervision: RG, WP, MIA, PCM

## DATA AND MATERIALS

Illumina reads and *de novo* assemblies will be made available on publication from the European Nucleotide Archive (GenBank project PRJEB71107). Consensus sequences are also available on line at https://www.doi.org/10.6084/m9.figshare.24874557.

