## Supplementary materials for "A putative hepatitis B virus sequence motif associated with hepatocellular carcinoma in South African adults"

**Supplementary methods:**

**Suppl Methods 1: Clinical and laboratory evaluation**

HCC diagnosis was made by clinical teams, using a combination of clinical evaluation, serum AFP results, radiological examination (at least one of abdominal ultrasound scan (USS), contrast-enhanced computed tomography (CT), or magnetic resonance imaging (MRI)), and liver biopsy in some cases (n = 25). Hepatitis B surface antigen (HBsAg) was confirmed in all individuals, tested using the Murex HBsAg Version 3 immunoassay kit (DiaSorin). HBV e-antigen (HBeAg) status was determined using the ETI-EBK PLUS assay (DiaSorin). HBV DNA viral load (VL) was measured using a validated in-house method as previously described[[1]](https://paperpile.com/c/WTPgdH/4oLpQ). Individuals were offered HIV tests on routine clinical grounds, which were performed through local validated diagnostic pathways in their local centre.

**Suppl Methods 2: Sequencing methods**

We sequenced plasma samples based on sufficient volume (≥ 0.5 mL) and a minimum HBV DNA viral load VL ≥4.0 log_10_ IU/mL (the limit of sensitivity for current WGS approaches at the time of the study[[2,3]](https://paperpile.com/c/WTPgdH/cls7H+xYfKb)). We extracted total nucleic acid from 0.5 mL of plasma using the NucliSENS magnetic extraction system (BioMérieux) and eluted into 25 μL of nuclease-free water.

We adapted a previously published protocol, to optimise for a larger volume input [[4]](https://paperpile.com/c/WTPgdH/1iRfD). The single-stranded (ss)DNA region of the partially double-stranded (ds) genome was completed to generate a fully double-stranded molecule. We purified and concentrated the nucleic acid with Agencourt RNAClean XP magnetic beads (Beckman Coulter). Sequencing libraries were generated using the Nextera DNA Library Preparation Kit (Illumina), and the DNA library size was assessed with the TapeStation system (Agilent) and Qubit dsDNA HS Assay (Thermo Fisher Scientific). We pooled samples together on an equimolar basis after indexing, and grouped samples into low or high VL pools (<6log_10_ IU/ml and ≥6log_10_ IU/ml, respectively) for enrichment. We used a target-enrichment workflow modified from the SeqCap EZ (Roche) protocol, using custom-designed pangenotypic HBV probes from IDT (xGen Lockdown Probes) as previously described[[5]](https://paperpile.com/c/WTPgdH/OoOGY). Samples were sequenced on an Illumina Mi-Seq using a v3 (250bp paired-end).

**Suppl Methods 3: Analysis of sequence data**

**Viral genome assembly**

We demultiplexed the data using QUASR v7.01[[6]](https://paperpile.com/c/WTPgdH/j3rgs) and removed adapter sequences with CutAdapt v1.7.1[[7]](https://paperpile.com/c/WTPgdH/y1L0s). We discarded any reads <50bp in length or mapping to the human reference sequence using Bowtie v2.2.4[[8]](https://paperpile.com/c/WTPgdH/zSEsp). We mapped remaining reads to a set of HBV sequences representing genotypes (A-I) using “BWA mem” v0.7.10[[9]](https://paperpile.com/c/WTPgdH/uGwvy) to choose an appropriate reference and to select HBV reads. Reads that mapped to at least one HBV reference sequence were then mapped to the closest? reference sequence using “BWA mem”. Consensus sequences were aligned and examined using the Simmonics software package[[10]](https://paperpile.com/c/WTPgdH/Eb9o5).

**Phylogenetic analysis**

From 154 full-length South African sequences from Genbank that were downloaded in November 2021[[11]](https://paperpile.com/c/WTPgdH/OJxnR), we removed those with >1% pairwise similarity (to remove duplicate sequences from the same sample/individual), leaving 61 sequences (**Suppl. Table 1**). We also used non-recombinant genotype reference sequences [[12]](https://paperpile.com/c/WTPgdH/t3B7d) and consensus sequences from our Illumina dataset (genotypes listed in **Suppl Table 3**).

We performed nucleotide alignments with Clustal X 2.1[[13]](https://paperpile.com/c/WTPgdH/bHwHa), and undertook phylogenetic analysis using a Bayesian Markov Chain Monte Carlo (MCMC) approach as implemented in the Bayesian Evolutionary Analysis by Sampling Trees (BEAST) version 1.10.4 program[[14]](https://paperpile.com/c/WTPgdH/56FRW) with an uncorrelated log-normal relaxed molecular clock, general time-reversible model, and nucleotide site heterogeneity estimated using a gamma distribution. The MCMC analysis was run for a chain length of 500,000,000. and results were visualized to confirm adequate chain convergence with Tracer version 1.7.2. The effective sample size (ESS) was calculated for each parameter, and all ESS values were >200 indicating sufficient sampling. The maximum clade credibility tree was selected from the posterior tree distribution after a 10% burn-in using Tree Annotator version 1.10.4 and visualized in FigTree version 1.4.4 as described previously[[15,16]](https://paperpile.com/c/WTPgdH/fEjsP+tz1mc).

**Supplementary results:**

**Suppl results 1: Patient characteristics**

In the HCC group, median age was significantly younger in patients with genotype A (median 33 years, IQR 30-43) compared to genotypes D or E (median 50 years, IQR 49-51 years) (p = 0.033, Fisher’s exact test). In contrast, this age difference was not observed in patients without HCC.

**Suppl results 2:** **Analysis of minority variants**

Several regions of the HBV genome were enriched for deletions in the HCC group, including PreS2 and core (spanning nt 2000-2400) (Figure 4, Suppl Figure 2). In PreS2, none of the sites in the non-HCC group had a deletion frequency >20% in contrast with the HCC group in which deletions were consensus in 7 cases. In core, point deletions in >20% sequences were identified at nt 2085 in three sequences (n = 2 HCC and n =1 non-HCC). Elsewhere in core, three HCC sequences had deletions in >20% of reads, including a 3bp consensus deletion in one sequence at nt 2349-2351 (Figure 4).

Several sequences had minority variant deletions at nt 2085 (core), although there was no association with HCC status. This is a homopolymeric region, and frameshift deletions have been described previously, particularly associated with HIV coinfection in HBV genotype A[[17–19]](https://paperpile.com/c/WTPgdH/UZzP4+owbHN+MaQFp). These mutations may generate a truncated pre-core protein that induces retention in the ER[[18]](https://paperpile.com/c/WTPgdH/owbHN). However, Illumina sequencing is notoriously inaccurate around homopolymeric regions[[20]](https://paperpile.com/c/WTPgdH/uyjxk).

**Supplementary Tables:**

**Suppl Table 1: Characteristics of adults with chronic hepatitis B virus infection (CHB) recruited in South Africa, comparing individuals with and without HCC.** Characteristics of the group with HCC and the group without HCC[[21,22]](https://paperpile.com/c/WTPgdH/YB0E8+ZGvqT) were compared.

| **Characteristic** | **HCC**  **(n = 68)** | **Non-HCC**  **(n = 93)** | **p-value†** |
| --- | --- | --- | --- |
| Age in years, median  (IQR) | 41  (33 - 51) | 36  (30 - 46) | **0.01** |
| Male sex, n  (%) | 55  (80.9) | 43  (46.2) | **<0.0001** |
| Positive HBeAg status  (%) | 20/65*  (30.8) | 19/93  (20.4) | 0.14 |
| HBV DNA VL, log_10_ IU/mL, median  (IQR) | 5.2  (3.5 - 6.9) | 3.5  (2.5 - 4.7) | **<0.0001** |
| Proportion with HBV DNA VL >20 IU/mL (%) | 48/61*  (78.7) | 53/92  (57.6) | **0.009** |
| Positive HIV status  (%) | 18/65*  (27.8) | 46/93  (49.5) | **0.006** |
| Proportion of HIV positive patients on cART (%) | 11/14*  (78.6%) | 46/46  (100%) | **0.001** |

* Denominator modified according to data availability. †Continuous variables are compared using the Kruskal-Wallis Rank Sum Test. Categorical variables were compared using the Chi-squared test when there were ≥5 observations in each group, otherwise Fisher’s exact test was used. Significant results (p ≤ 0.05) appear in bold. IQR - interquartile range; cART - combined antiretroviral therapy; HBeAg - Hepatitis B virus e-antigen; VL - viral load.

**Suppl Table 2:** **Characteristics of cohorts and output of deep sequencing by Illumina for HBV infection, comparing individuals with and without HCC.** The 33 subjects presented here are a subset of those presented in **Suppl Table 1**, summarising samples with viral loads (VL) ≥4.0 log_10_ IU/ml with sufficient sample (≥0.5 mL plasma) for deep sequencing.

|  | **HCC**  **(n = 19)** | **Non-HCC**  **(n = 14)** | **p-value†** |
| --- | --- | --- | --- |
| Age in years, median  (IQR) | 35.0  (30.2, 47.8) | 40.0  (36.0, 45.0) | 0.482 |
| Male sex  (%) | 16 / 19  (84.2) | 9 / 14  (64.3) | 0.238 |
| Positive HBeAg status  (%) | 9 / 19  (47.4) | 8 / 14  (57.1) | 0.839 |
| HBV DNA VL, log _10_ IU/mL, median  (IQR) | 6.2  (5.5, 7.6) | 4.9  (4.7, 5.5) | **0.015** |
| Positive HIV status  (%) | 7 / 19  (36.8) | 6 / 14  (42.9) | 1 |
| Genotype A (%) | 15 / 19  (78.9) | 10 / 14  (71.4) | 0.695 |
| Genotype D (%) | 2 / 19  (10.5) | 3 / 14  (21.4) | 0.628 |
| Genotype E (%) | 2 / 19  (10.5) | 1 / 14  (7.1) | 1 |
| Median HBV genome coverage, reads/site after deduplication* (IQR) | 1,121  (117 – ~~-~~ 4,263) | 177  (37 – ~~-~~ 358.3) | 0.13 |
| Proportion of reads mapping to HBV (IQR) | 28.5%  (5.3 – ~~-~~ 41.3) | 9.6%  (3.6 – ~~-~~ 37.9) | 0.48 |

†Continuous variables are compared using Kruskal-Wallis Rank Sum Test. Categorical variables were compared using the Chi-Square test where there were ≥5 observations in each group, otherwise Fisher’s exact test was used. Significant results (p ≤ 0.05) appear in bold. The median number of total reads generated per sample was 442,748 (IQR 242,566-1,032,070).

**Suppl Table 3: Number of sequences by HBV genotype**

| **Genotype** | **New South Africa cohort reported in this paper (% of total)** | **Full length sequences published from South Africa* (% of total)** |
| --- | --- | --- |
| A1 | 22 (71.0) | 42 (68.9) |
| A2 | 1 (3.2) | 7 (11.5) |
| B | 0 | 1 (1.6) |
| C | 0 | 3 (4.9) |
| D | 5 (16.1) | 6 (9.8) |
| E | 3 (9.7) | 1 (1.6) |
| G | 0 | 1 (1.6) |
| **TOTAL** | **31** | **61** |

***Accession numbers of 61 South African sequences downloaded from GenBank**

AY233289.1, AY233287.1, AY233278.1, KU605532.1, AY233288.1, AF297624.1, AY233274.1, KM519453.1, AY233284.1, GQ184322.1, AY233279.1, AY233286.1, KT347090.1, MT210032.1, HQ646553.1, KF922409.1, GQ184326.1, AY233281.1, KF922431.1, AY903452.1, JN182333.1, GQ184325.1, KU605533.1, HQ646556.1, AY233296.1, HQ646552.1, AF297620.1, KT347092.1, KF922423.1, AF297625.1, KF922437.1, AF297621.1, AY233277.1, MT210034.1, KF922421.1, AF297622.1, KF922432.1, U87742.3, AF297623.1, MH481862.1, AY233290.1, U87746.3, U87747.3, KY004111.1, KF922413.1, KT347088.1, AY233276.1, KF922425.1, KP234050.1, KF922428.1, KT347087.1, GQ167302.1, AY233294.1, KU605539.1, KF922439.1, HQ646555.1, KJ010778.1, KT347089.1, KF922430.1, AF297619.1, and KF922434.1

**Supplementary Figures:**

**Suppl Figure 1: Schematic showing number of samples in each phase of the study, with (i) adults recruited through clinical studies in South Africa, and (ii) downloaded South African sequence data from GenBank.** (A) 161 adults with HBsAg infection were characterised at baseline in HCC and non-HCC groups (Table 1), (B) 33 samples with HBV DNA that met the threshold for Illumina sequencing, (C) 31 samples for which WGS assembly was achieved.

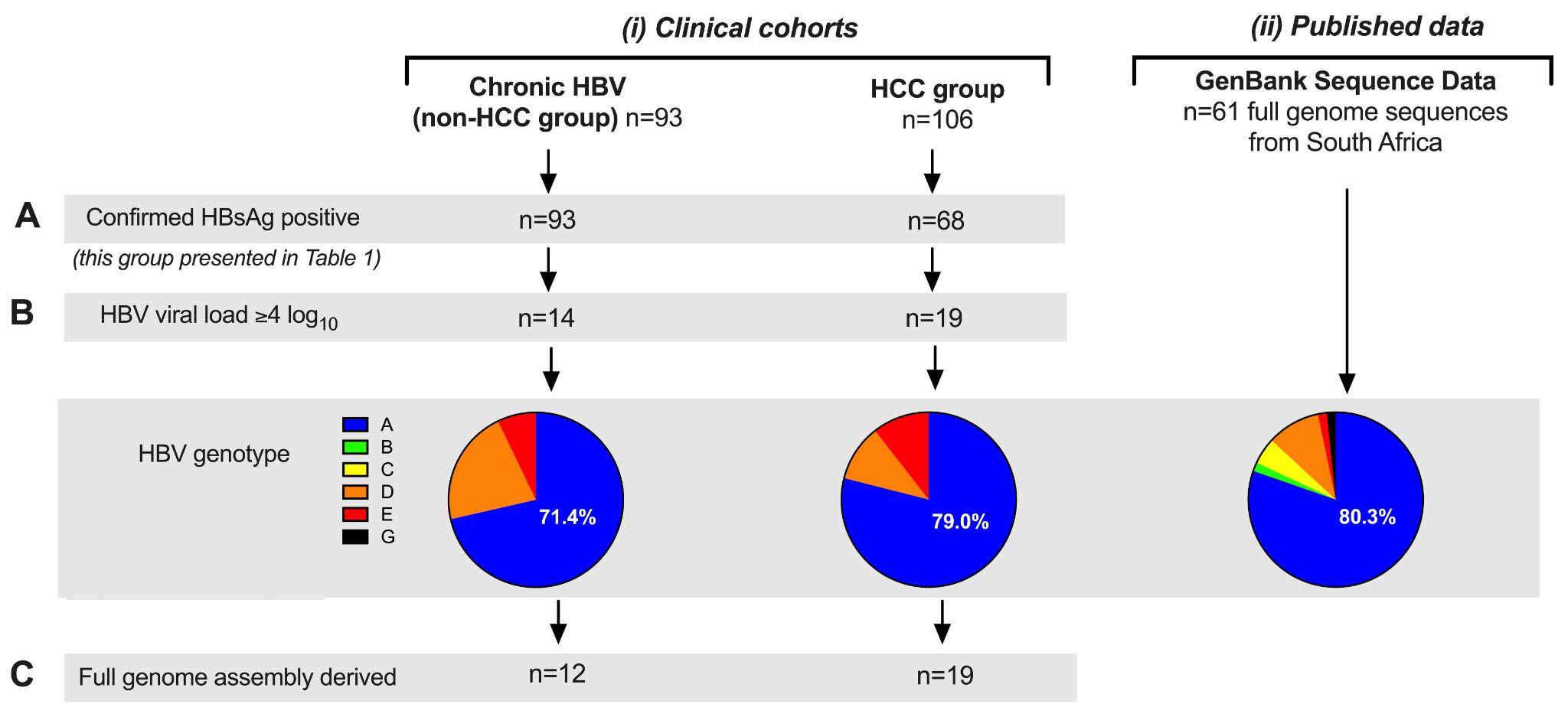

**Suppl Figure 2. HBV DNA Viral load (VL), number of templates sequenced, and read coverage of the HBV genome from Illumina sequencing** (A) HBV DNA VL from plasma samples from n = 33 patients with hepatocellular carcinoma (19 HCC) and without (14 non-HCC). Median values and the inter-quartile ranges (IQR) are indicated. Note that only samples with VL ≥4.0 log_10_ IU/mL underwent Illumina sequencing. (B) Unique (deduplicated) HBV templates recovered through Illumina sequencing for the HCC (n = 19) and non-HCC (n = 14) samples. (C) Median and IQR coverage (reads that span each site, also known as “read depth”) of the HBV genome for the HCC (n = 19) and non-HCC (n = 12) samples for which non-zero *de novo* assemblies could be inferred. Nucleotide positions are with respect to the HBV accession X02763 (genotype A) reference strain. Note that in (A,B), values for samples that generated non-zero length *de novo* assemblies denoted in (C) are indicated by triangles; the n = 2 non-HCC samples that could not be assembled appear as circles.

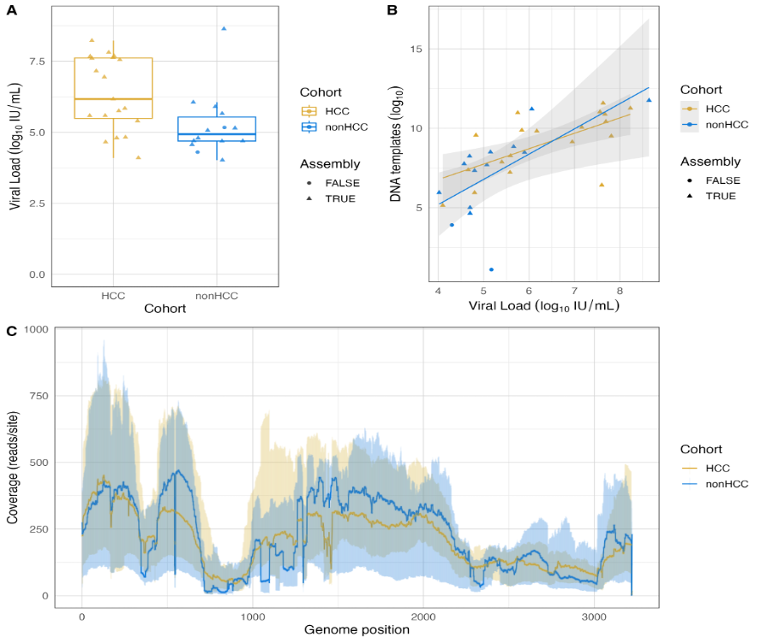

**Suppl Figure 3. Phylogenetic tree of HBV sequences from South African adults with (HCC) and without (non-HCC) hepatocellular carcinoma, together with other published full-length sequences from South Africa and genotype reference sequences.** Tree constructed using a Bayesian Markov Chain Monte Carlo (MCMC) approach with an uncorrelated log-normal relaxed molecular clock, general time-reversible model, and nucleotide site heterogeneity estimated using a gamma distribution. Sequences from the HCC group are shown in blue (n = 19), non-HCC in green (n = 12), reference sequences of known genotype in black (n = 49) and published full length sequences from South Africa downloaded from GenBank shown in red (n = 61) see **Suppl. Table 3** for list of sequences. Posterior probabilities are indicated at the nodes, with values >0.90 typically considered significant. Note that GenBank metadata for KM519454 has labeled the sequence as gtA1, but the sequence clusters with gtA2 sequences suggesting the assigned subgenotype is incorrect.

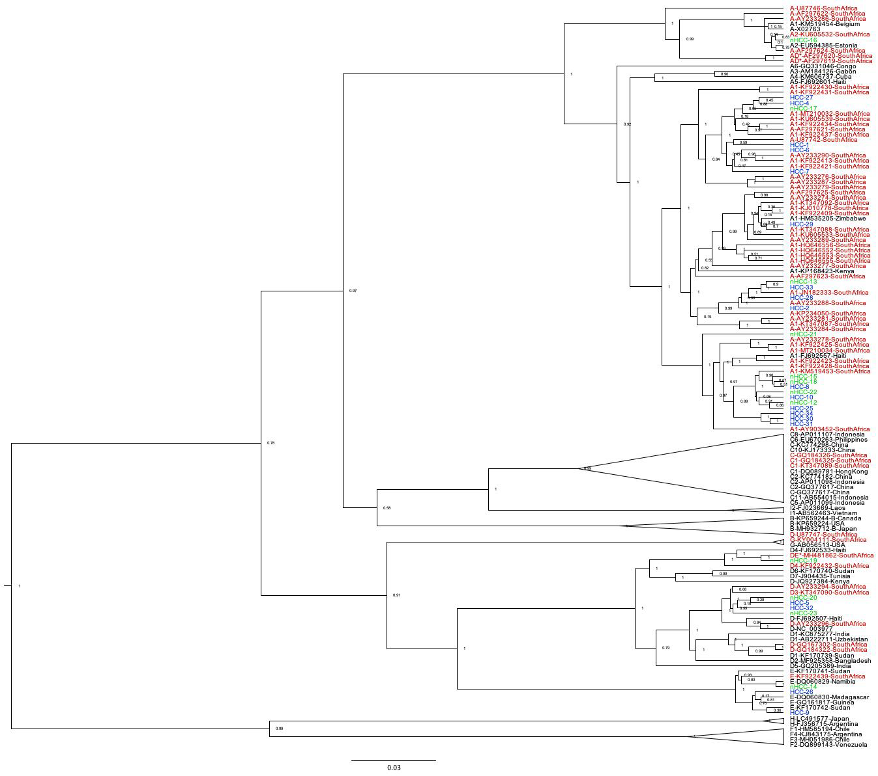
